# Smoking and the risk of COVID-19 in a large observational population study

**DOI:** 10.1101/2020.06.01.20118877

**Authors:** Ariel Israel, Elan Feldhamer, Amnon Lahad, Diane Levin-Zamir, Gil Lavie

## Abstract

**BACKGROUND:** Smokers are generally more susceptible to infectious respiratory diseases and are at higher risk of developing severe complications from these infections. Conflicting reports exist regarding the impact of smoking on the risk of Coronavirus disease 2019 (COVID-19).

**METHODS:** We carried out a population-based study among over 3,000,000 adult members of Clalit Health Services, the largest health provider in Israel. Since the beginning of the disease outbreak, and until May 16, 2020, over 145,000 adults underwent RT-PCR testing for Severe Acute Respiratory Syndrome Coronavirus 2 (SARS-CoV-2), and 3.3% had positive results. We performed a case-control study among patients who underwent SARS-CoV-2 testing, to assess the impact of smoking on infection incidence and severity. Individuals with positive tests were matched in a 1:5 ratio to individuals tested negative, of the same sex, age, and ethnicity/religion. Conditional logistic regressions were performed to evaluate odds ratios for current and previous smoking on the risk of testing positive. Multivariable logistic regressions were performed among patients infected with COVID-19 to estimate the association between smoking and fatal or severe disease requiring ventilation. Regressions were performed with and without adjustment for preexisting medical conditions.

**RESULTS:** Current smokers (9.8%) were significantly less prevalent among members tested positive compared to the general population (19.4%, P<0.001), and to matched members tested negative (18.2%, P<0.001). Current smoking was associated with significantly reduced odds ratio (OR) for testing positive OR=0.447 (95% confidence interval (CI) 0.400-0.501). Among patients tested positive, there was no evidence of significantly increased risk of developing severe or fatal disease.

**CONCLUSION:** The risk of infection by COVID-19 appears to be reduced by half among current smokers. This intriguing finding may reveal unique infection mechanisms present for COVID-19 which may be targeted to combat the disease and reduce its infection rate.

---

SARS-Cov-2 is a new virus, which was first identified in December 2019, and has rapidly spread into a global pandemic of primarily respiratory illness designated as Coronavirus Disease 2019 (COVID-19). This disease is associated with significant mortality, particularly among the aging population, raising considerable concerns for public health. Smokers are at greater risk for respiratory infections and were expected to be at increased risk for Covid-19 as well^1-3^. Nevertheless, some reports appear to indicate that SARS-Cov-2 infection rate is not higher among smokers, and in several cohorts, the infection rate among smokers was lower than in the general population^4-7^. There are also conflicting data regarding clinical outcomes for smokers, and whether smoking is a risk factor for more severe disease. A recent review published by the World Health Organization summarizes the conflicting results of the studies available to date, and the lack of knowledge regarding the association between smoking and the risk of SARS-CoV-2 infection and disease severity^8^, highlighting the need for population-based studies to better understand the association. We report here the results of a large population study performed among adult members of Clalit Health Services (CHS), the largest healthcare provider in Israel.

## Methods

### Study population and data collection

Clalit Health Services (CHS) provides comprehensive health services to over 3,000,000 adult members, and centrally manages electronic health records (EHR) including longitudinal records for over two decades. We anonymously collected selected variables from the EHR of patients who underwent SARS-Cov-2 testing and had documented smoking status. Demographic variables, including age, gender, religious/ethnic group, were collected from the CHS database. Preexisting conditions were considered present when a corresponding documented diagnosis was present in the EHR. Patients were considered obese if their last documented body mass index (BMI) was above 30, or if they had a documented diagnosis of obesity. Follow-up, including hospitalization, condition severity, the need for ventilation, and mortality were extracted from hospitalization records collected by the Israeli Ministry of Health, up to June 3, 2020.

CHS institutional review board approved the project with a waiver of informed consent, approval number: COM-0046-20.

### Statistical analysis

We identified adult patients, who underwent RT-PCR testing for SARS-CoV-2 until the cut-off date of May 16, 2020, and who aged, at the time of the test between 18 and 95 years. Within this population, we defined a matched cohort where each patient positive for SARS-CoV-2 was matched to five patients tested negative of the same gender, age group (in five years intervals), and religious/ethnic group. Conditional logistic-regression models were fitted for estimating the odds-ratio (OR) and corresponding 95% confidence interval (CI) for the risk of testing positive. A separate model, accounting for preexisting conditions, including chronic obstructive pulmonary disease (COPD), asthma, hypertension, obesity, reported arrhythmia, peripheral vascular disease, ischemic heart disease, and malignancy, was fitted to account for conditions potentially influencing the risk associated to smoking.

Among patients tested positive, we assessed the effect of smoking status on the risk of fatal or severe disease requiring mechanical ventilation, by fitting multivariable logistic models accounting for gender, age category, and religious/ethnic groups. A separate model was also performed to adjust for preexisting conditions.

Statistical significance of differences observed between groups was assessed by the Chi-Square test for categorical variables, and two tailed T-test for continuous variables. P-values below 0.05 were considered significant. Statistical analyses were performed using R statistical software version 3.6 (R Foundation for statistical computing).

### Results

The flowchart of the cohort creation is presented as Figure 1. From the beginning of the outbreak and until May 16, 2020,128,427 distinct CHS members with documented smoking status, underwent RT-PCR tests for SARS-CoV-2, 4,235 (3.3%) of whom tested positive. Among them, we found that 9.8% were current smokers and 11.7% past smokers, while their corresponding rates within the adult population of CHS were 19.4% and 13.9% respectively (P<0.001).

**Figure 1:**
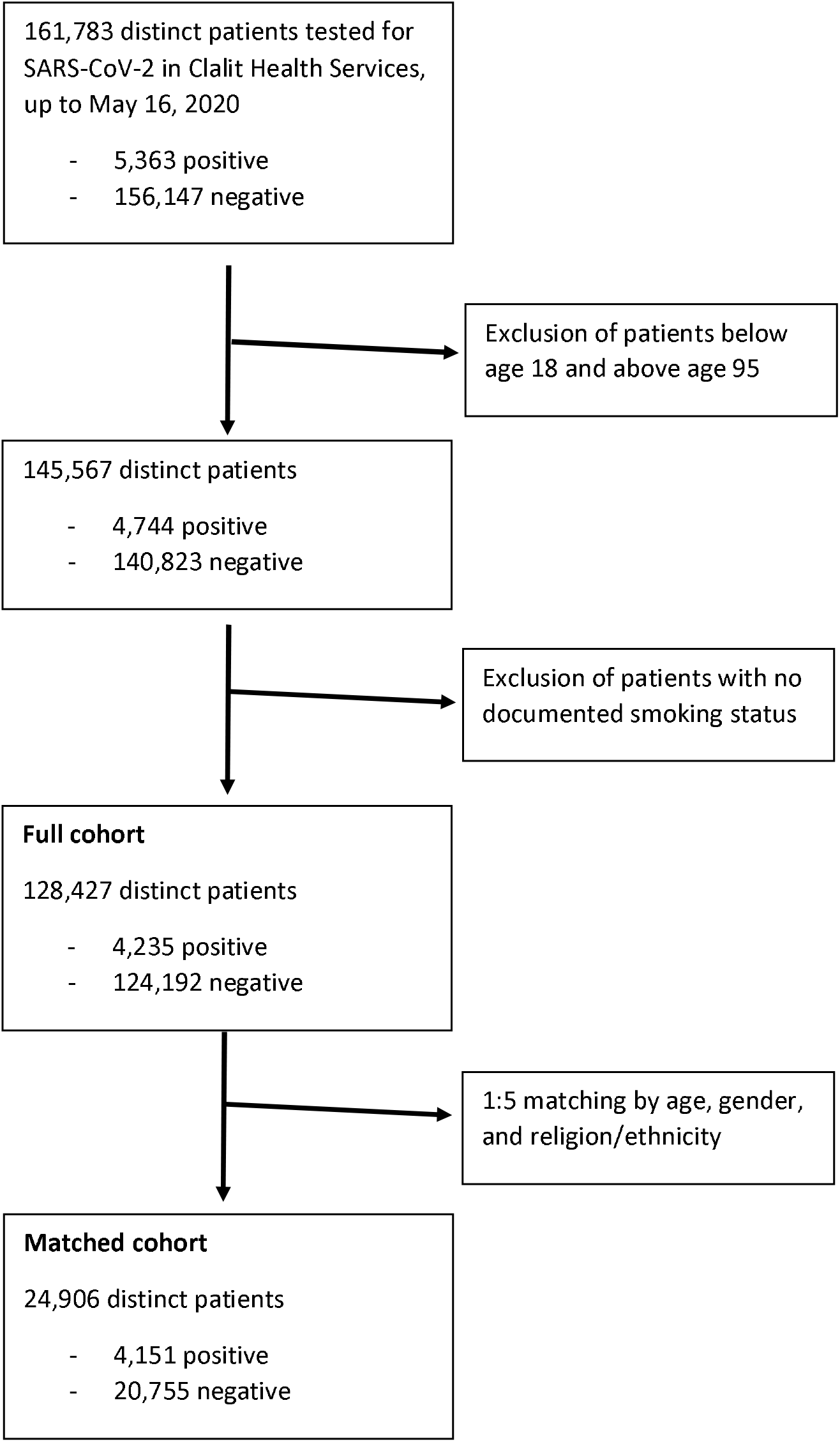
Cohort creation flowchart.

The characteristics of the matched cohort are shown in Table 1. The observed difference in smoking status is maintained in this cohort: current smoking prevalence is significantly lower among patients who tested positive (9.8%) than among matched patients tested negative (18.2%). Table 2 presents the regression results for testing positive for COVID-19 in the matched cohort. In column A, the results of the conditional logistic regression are presented, adjusted only for age, gender, and religion/ethnicity categories. In this model, current smoking was associated with significantly reduced OR 0.447, with a 95 % Cl of 0.400-0.501. Interestingly, odds for testing positive were also significantly reduced in past smokers, although the effect was more moderate: OR 0.757 (95% CI 0.679-0.844). We assessed these associations in a separate model adjusted for preexisting conditions and found very similar results for smoking (column B). Interestingly, odds for positive testing was increased for obese patients OR 1.172 (95% Cl 1.084-1.267) but decreased in hypertensive patients OR 0.834 (95% Cl 0.738-0.941), suggesting that among these two frequently correlated conditions, obesity is the one associated with increased infection rate. Odds ratio for infection were also decreased for patients with asthma or malignancy, likely reflecting increased adherence to social distancing recommendations among these patients.

**Table 1:** Demographics and Clinical characteristics of the matched cohort of patients who tested positive and negative for COVID-19.

|  | COVID-19 Positive<br>(Cases) | COVID-19 Negative<br>(Controls) |
| --- | --- | --- |
| N | 4,151 | 20,755 |
| Age (median [IQR]) | 39.54 [27.38,58.26] | 39.65 [27.55,58.67] |
| Gender female (%) | 1998 (48.1) | 9990 (48.1) |
| Smoking status (%) |  |  |
| never smoker | 3,262 (78.5) | 14,301 (68.9) |
| **current smoker** | **406 (9.8)** | **3,783 (18.2)** |
| **past smoker** | **483 (11.6)** | **2,671 (12.9)** |
| Ethnicity/Religion (%) |  |  |
| Jewish/General | 2,466 (59.4) | 12,330 (59.4) |
| Arab | 672 (16.2) | 3,360 (16.2) |
| Jewish Orthodox | 1,013 (24.4) | 5,065 (24.4) |
| Age group (%) |  |  |
| 18-34 | 1,748 (42.1) | 8,740 (42.1) |
| 35-54 | 1,161 (28.0) | 5,805 (28.0) |
| 55-74 | 998 (24.0) | 4,990 (24.0) |
| 75- | 244 (5.9) | 1,220 (5.9) |
| Body Mass Index (BMI) (mean (SD)) | 27.35 (5.74) | 26.99 (5.73) |
| BMI group (%) |  |  |
| <20 | 211 (5.1) | 1,216 (5.9) |
| 20-25 | 997 (24.0) | 5,331 (25.7) |
| 25-30 | 1,031 (24.8) | 5,322 (25.6) |
| 30- | 909 (21.9) | 4,059 (19.6) |
| missing BMI | 1,003 (24.2) | 4,827 (23.3) |
| Number of visits to primary doctor (mean (SD)) in the last year | 4.88 (5.37) | 5.46 (6.43) |
| **Chronic conditions:** |  |  |
| Chronic obstructive pulmonary disease (COPD) | 80 (1.9) | 599 (2.9) |
| Asthma | 195 (4.7) | 1286 (6.2) |
| Hypertension | 655 (15.8) | 3,599 (17.3) |
| Obesity * | 1269 (30.6) | 5,895 (28.4) |
| Peripheral vascular disease | 50 (1.2) | 353 (1.7) |
| Ischemic Heart Disease | 236 (5.7) | 1,371 (6.6) |
| Malignancy | 231 (5.6) | 1,438 (6.9) |
| **COVID-19 hospitalization status (%) <sup>1</sup>** |  |  |
| hospitalized in good condition | 623 (15.0) |  |
| hospitalized in moderate condition | 166 (4.0) |  |
| hospitalized in severe condition | 85 (2.0) |  |
| deceased | 75 (1.8) |  |

IQR: Interquartile range
\* In this and other tables, obesity is defined as presence of obesity diagnosis in the electronic medical record or last measured body mass index (BMI) above 30. <sup>1</sup> Worst condition of the patient
during hospitalization: patients who deceased were not counted as hospitalized in good, moderate or severe condition.

**Table 2:** Conditional logistic regression for estimating smoking and comorbidity effects on SARS-CoV-2 infection status in the matched cohort. *(N = 24,906; positive outcome: 4,151 who had positive test)*

|  | **(A) Conditional** |  |  | **(B) Adjusted for comorbidity** |  |  |
| --- | --- | --- | --- | --- | --- | --- |
|  | OR | CI (95%) | p-value | OR | CI (95%) | p-value |
| **Smoking status:** |  |  |  |  |  |  |
| never smoker |  | reference |  |  | reference |  |
| **current smoker** | 0.447 | 0.400-0.501 | <0.001 | 0.453 | 0.405-0.507 | <0.001 |
| past smoker | 0.757 | 0.679-0.844 | <0.001 | 0.773 | 0.693-0.863 | <0.001 |
| **Chronic conditions:** |  |  |  |  |  |  |
| Chronic obstructive pulmonary disease (COPD) |  |  |  | 0.880 | 0.687-1.128 | 0.314 |
| Asthma |  |  |  | 0.766 | 0.654-0.896 | 0.001 |
| Hypertension |  |  |  | 0.834 | 0.738-0.941 | 0.003 |
| Obesity |  |  |  | 1.172 | 1.084-1.267 | <0.001 |
| Arrhythmia |  |  |  | 0.906 | 0.756-1.085 | 0.285 |
| Peripheral vascular disease |  |  |  | 0.839 | 0.615-1.146 | 0.271 |
| Ischemic Heart Disease |  |  |  | 0.936 | 0.791-1.108 | 0.445 |
| Malignancy |  |  |  | 0.788 | 0.677-0.917 | 0.002 |

Model (A) is based on a conditional logistic regression adjusted for age, gender and ethnicity/religion strata. Model (B) is based on multivariable logistic regression accounting for age, gender and ethnicity strata, and the assessed chronic conditions (COPD, asthma, hypertension, obesity, reported arrhythmia, peripheral vascular disease, ischemic heart disease, and malignancy)

Table 3 describes the odds ratio for the risk of severe or fatal disease among patients positive for SARS-CoV-2 in the cohort. Severe or fatal disease was very strongly associated with increasing age and with male gender, as evidenced by most studies to date. However, we found no statistically significant association between current or past smoking and disease severity, both in the model adjusted for baseline demographic characteristics (model A, adjusted for age, gender, and religious/ethnic group), and in the model that accounted for preexisting conditions as well (model B); in the latter model, presence of ischemic heart disease or malignancy was associated with significantly increased odds ratio for severe or fatal disease.

**Table 3:** Logistic regression for estimating smoking, demographic and comorbidity effects on fatal or severe disease among SARS-CoV-2 positive. *(N = 4,151; positive outcome: 160 who had severe or fatal disease)*

|  | (A) Adjusted for age, sex, and ethnic/religious sector |  |  | (B) Adjusted for comorbidity |  |  |
| --- | --- | --- | --- | --- | --- | --- |
|  | OR | CI (95%) | p-value | OR | CI (95%) | p-value |
| **Smoking status:** |  |  |  |  |  |  |
| never smoker |  | reference |  |  | reference |  |
| **current smoker** | 0.618 | 0.289-1.323 | 0.215 | 0.586 | 0.265-1.296 | 0.187 |
| past smoker | 1.347 | 0.903-2.010 | 0.144 | 1.108 | 0.729-1.687 | 0.631 |
| Gender female | 0.620 | 0.435-0.883 | 0.008 | 0.644 | 0.444-0.933 | 0.020 |
| **Age group:** |  |  |  |  |  |  |
| 18-34 |  | reference |  |  | reference |  |
| 35-54 | 3.486 | 1.334-9.114 | 0.011 | 3.118 | 1.189-8.178 | 0.021 |
| 55-74 | 23.873 | 10.275-55.468 | <0.001 | 14.992 | 6.253-35.944 | <0.001 |
| 75+ | 90.593 | 38.102-215.399 | <0.001 | 39.321 | 15.414-100.311 | <0.001 |
| **Ethnic/religious sector:** |  |  |  |  |  |  |
| General (Jewish) |  | reference |  |  | reference |  |
| Arab | 0.970 | 0.535-1.758 | 0.920 | 0.926 | 0.502-1.708 | 0.805 |
| Jewish Orthodox | 0.999 | 0.656-1.522 | 0.997 | 1.111 | 0.722-1.710 | 0.631 |
| **Chronic conditions:** |  |  |  |  |  |  |
| Chronic obstructive pulmonary disease (COPD) |  |  |  | 0.987 | 0.485-2.011 | 0.972 |
| Asthma |  |  |  | 0.986 | 0.471-2.066 | 0.971 |
| Hypertension |  |  |  | 1.252 | 0.826-1.899 | 0.290 |
| Obesity |  |  |  | 1.131 | 0.788-1.624 | 0.503 |
| Arrhythmia |  |  |  | 1.342 | 0.828-2.175 | 0.233 |
| Peripheral vascular disease |  |  |  | 1.606 | 0.771-3.347 | 0.206 |
| Ischemic Heart Disease |  |  |  | 2.023 | 1.299-3.149 | 0.002 |
| Malignancy |  |  |  | 3.071 | 2.018-4.672 | <0.001 |

Model (A) is based on a multivariable logistic regression adjusted for age, gender and ethnic/religious sector. Model (B) is based on multivariable logistic regression accounting for age, gender, ethnicity, and the assessed chronic conditions (COPD, asthma, hypertension, obesity, reported arrhythmia, peripheral vascular disease, ischemic heart disease, and malignancy)

### Discussion

In this large population of patients tested for COVID-19 infection, there appears to be significantly reduced risk for COVID-19 infection among smokers, in contrast to what occurs in most respiratory infections. This intriguing observation confirms several recently published studies, that also reported decreased disease incidence among smokers^4,6,7,9^, although in a smaller scale. Interestingly, even among SARS-CoV-2 positive patients, we found no evidence of significant association between current or past smoking and disease severity, as reflected by death during hospitalization or by need for mechanical ventilation.

To our knowledge, this is the first study to assess the association between smoking and COVID-19 infection in a matched cohort of this scale. The magnitude of association observed for current smoking, with odds of infection reduced by about a half in smokers, suggests a genuine protective effect of smoking on the risk of COVID-19. A significant negative association is also observed between past smoking and SARS-CoV-2 infection, although more moderate than the one observed for current smoking. It is unclear whether the latter association reflects long-term changes engendered by past smoking, or unreported smoking recidivism among these patients. Among infected patients, we did not observe a significant association between current smoking and severity of disease, in contrast to what occurs in most respiratory infections. Other national cohorts did not observe an increased risk for severe disease among smokers as well, while some smaller studies sampling hospitalized patients observed increased risk among smokers^1,2,10^. These apparently conflicting results may reflect different thresholds for patient inclusion. Our cohort included many patients who were asymptomatic or had light symptoms and did not undergo hospitalization. It is possible that among study samples based on patients hospitalized for serious disease, smoking or past smoking contributes to disease aggravation. Future studies should examine more in-depth specific smoking habits, for example light to heavy smoking, to discern whether there is a possible dose response, or whether the possible protective effect is based on a specific threshold.

The negative association observed here between smoking and COVID-19 infection incidence is uncommon among respiratory infections, and possibly reflects unique infection mechanisms present in the novel coronavirus. Changeux et a I^11^, relying on similar observations, propose a crucial role for the nicotinic acetylcholine receptor (nAChR) in COVID-19 pathology. According to their neurotropic hypothesis, SARS-CoV-2 invades the central nervous system through the nAChR receptor, present in neurons of the olfactory system, as reflected by the frequent occurrence of neurologic symptoms, such as loss of smell or taste, or intense fatigue in patients affected by COVID-19. Other mechanisms may also affect SARS-CoV-2 infection potential in smokers. It is widely accepted that the angiotensin converting enzyme 2 (ACE2) represents the main receptor molecule for SARS-CoV-2, and smoking has been shown to differentially affect ACE2 expression in tissues^12-14^. Other putative explanations could involve altered cytokine expression such as IL-6, for which increased levels are associated with unfavorable disease outcome^14,15^.

The effect of smoking among the study population was assessed through self-reported information routinely collected during medical visits, prior to occurrence of COVID-19, thus eliminating possible recall bias by patients. The matched cohort design, and the decision to only include patients that have been tested for SARS-CoV-2, overcomes the potential confounding effects of age, gender, and ethnicity/religious group, on smoking habits. We acknowledge our study’s limitations as observational, noting the difficulty in eliminating all possible confounders. However, in the context of a harmful and addictive habit such as smoking, a controlled trial is not ethical.

Acknowledging the destructive effects of smoking on health, the importance of smoking prevention and cessation to preserve health, and the highly addictive nature of nicotine, we strongly encourage all patients to refrain from smoking, as the long term effects of this hazardous habit far outweigh potential benefits in preventing SARS-CoV-2 infection. Nevertheless, the strong negative association demonstrated in this study between smoking and COVID-19 incidence may offer promising new directions for fighting this disease, based on a better understanding of the mechanisms by which smoking reduces the risk of SARS-CoV-2 infection.

## Data Availability

Not applicable.

## Acknowledgement

All authors have no conflict of interest to report

No external funding was received for this study

## Notes

### Competing Interest Statement

The authors have declared no competing interest.

### Author Declarations

Clalit Health Services Institutional Review Board

